# Cohort profile: KaroLiver, a population-based cohort of patients receiving curative liver-directed treatment for colorectal liver metastases at a Swedish tertiary centre

**DOI:** 10.64898/2026.08.24.26361235

**Authors:** Marco Gerling, Carlos Fernández Moro, Celina Limbecker, Annika Viljamaa, Sara Harrizi, Yousra Hamidi, Ann-Kathrin Hailer, Johanna Sterner, Ernesto Sparrelid, Lorand Bozóky, Evelina Tidholm Qvist, Ruth Baumgartner, Media Salmonson Schaad, Béla Bozóky, Natalie Geyer, Jennie Engstrand

**Affiliations:** Department of Clinical Science, Intervention and Technology, Division of Surgery and Oncology, Karolinska Institutet, Stockholm, Sweden; Theme Cancer, Karolinska University Hospital, Stockholm, Sweden; Department of Laboratory Medicine, Division of Pathology, Karolinska Institutet, Stockholm, Sweden; Department of Clinical Pathology and Cancer Diagnostics, Karolinska University Hospital, Stockholm, Sweden

## Abstract

**Purpose:** The Karolinska Liver Metastases (KaroLiver) cohort was established to investigate associations between clinical characteristics and histopathological features in patients treated with curative intent for colorectal cancer liver metastases (CRLM). The cohort combines whole-slide digital histopathology images with detailed oncological, surgical, radiological and survival data, enabling comprehensive analyses of treatment trajectories, clinical outcomes and metastatic tumour biology.

**Participants:** KaroLiver is a retrospective observational cohort comprising all consecutive patients who received curative-intent, liver-directed treatment for CRLM at Karolinska University Hospital in Stockholm, Sweden. The hospital is the primary regional referral centre for HPB surgery, serving the population of approximately 2.5 million people in the Stockholm-Gotland healthcare region. Patient enrolment is continuously updated in accordance with amended ethical approvals and evolving scientific questions. The cohort currently comprises 811 patients who underwent 1204 liver interventions between February 2012 and January 2022. Detailed clinical, oncological, surgical, pathological, molecular, recurrence and survival data are collected.

**Findings to date:** Median overall survival (OS) in the current cohort is 51.0 months (95% CI 46.2–57.1 months), and median recurrence-free survival (RFS) is 10.4 months (95% CI 9.2–11.9 months). The five-year OS rate is 44.9% (95% CI 41.2–48.8%). Studies using the cohort have so far identified a liver injury-derived stromal capsule in a subset of metastases, associated with improved survival. The cohort has also enabled the identification of histopathological markers of tumour biology, sex-based differences in treatment and survival, and associations between post-hepatectomy liver failure and oncological outcomes.

**Future plans:** Current research priorities include advanced histology-based prognostic scoring, sex differences in recurrence and retreatment, tumour biology and outcomes in early-onset versus average-onset CRLM, as well as CT- and MRI-based radiomics, all integrated within KaroLiver’s histopathological framework. Data sharing is supported, given that regulatory requirements are met. Retrospective accrual and outcome updates will continue for current and future studies, subject to the required approvals.

**Strengths and limitations of this study:**

□ Selection bias in KaroLiver is minimised through consecutive, population-based accrual of all patients receiving curative-intent liver-directed treatment for CRLM at a high-volume Swedish tertiary centre.
□ Granular data on recurrence, including site, timing, and intent of retreatment, enable detailed analyses of disease trajectories beyond the index hepatectomy.
□ KaroLiver is linked to CT and MRI examinations from diagnosis through recurrence; combined with detailed histopathological and molecular pathology data, this provides a platform for translational research, based on specific, defined research questions.
□ The observational design limits causal inference, and changes in treatment strategies and follow-up protocols over the study period may introduce temporal confounding that must be accounted for in analyses.
□ Molecular data, including *RAS*/*RAF* mutation status and mismatch repair status, are incomplete, reflecting real-world clinical practice and the gradual adoption of tumour profiling over the study period.

## Introduction

Colorectal cancer (CRC) is one of the most common malignancies globally. Approximately 25% of patients with CRC develop colorectal liver metastases (CRLM) during the course of their disease [1]. Surgical resection or ablation remains the cornerstone of potentially curative treatment, with five-year survival rates of 40–55% among selected patients [1]. Despite advances in systemic therapy, surgical techniques, and perioperative care, tumour recurrence is common, affecting more than half of all patients within three years of curative-intent treatment [2,3].

Over the past two decades, innovations in thermal ablation, staged hepatectomy, and parenchyma-sparing techniques have expanded eligibility for liver-directed treatment to patients with greater metastatic burden, bilobar disease, or limited extrahepatic disease [2,4]. This broadening of treatment eligibility, alongside improved systemic therapies, has increased the clinical heterogeneity of patients undergoing liver-directed therapy and reinforced the need for improved prognostic and treatment-selection tools.

CRLM is increasingly recognised as a biologically and clinically heterogeneous disease, with outcomes shaped by both tumour biology and the extent and distribution of metastatic disease. Histopathological features such as growth patterns and resection-margin status, together with molecular characteristics including *RAS*/*RAF* mutations and mismatch repair status, influence recurrence risk, treatment decisions, and survival [5–7]. However, these factors remain incompletely integrated into routine clinical decision-making.

Histological growth patterns are of particular interest for understanding metastasis biology as they reflect distinct modes of interaction between metastatic tumour cells and the surrounding liver parenchyma that shape outcomes. These patterns provide prognostic information independent of established clinical and pathological factors, yet their biological determinants and potential therapeutic implications remain incompletely understood [6,7], and they are not yet integrated into clinical decision algorithms.

Radiologically assessed tumour burden and distribution also strongly influence prognosis, and emerging radiomics approaches hold promise for the non-invasive prediction of outcomes [8], although their clinical utility requires validation in large, well-characterised cohorts.

Recurrence does not necessarily preclude further curative-intent treatment, as repeat liver intervention can provide substantial survival benefit in selected patients [9]. Detailed characterisation of the site and timing of recurrence, as well as eligibility for and intent of retreatment, is therefore essential for understanding the full disease trajectory. Although early detection of recurrence is considered important, the optimal intensity and structure of postoperative follow-up remain controversial [10].

Patient-related factors further contribute to variations in outcomes. For example, sex-based differences in CRC and CRLM incidence, metastatic patterns, access to surgery, and postoperative outcomes have been reported, although more research is needed to clearly define how sex impacts on CRLM diagnosis and treatment [11,12]. Similarly, early-onset CRC, defined as CRC diagnosed before the age of 50 years, is increasing in incidence and may represent a biologically distinct entity, often presenting with advanced or metastatic disease [13,14]. However, whether sex and age are associated with distinct patterns of metastatic spread, resectability, recurrence, or access to retreatment remains insufficiently studied. Other patient characteristics, including comorbidity and frailty, may also increasingly influence treatment selection as eligibility for liver-directed therapy expands.

Against this background, clinically important questions remain regarding how CRLM biology, disease distribution, treatment characteristics, and patient-related factors jointly shape recurrence and long-term outcomes. The KaroLiver cohort was established to enable comprehensive clinical, translational, and health services research across the full disease trajectory of CRLM. A defining feature of the cohort is the systematic linkage of detailed clinical, surgical, radiological, pathological, recurrence and survival data with whole-slide digital histopathology. By integrating these data, KaroLiver is positioned to address questions spanning treatment outcomes, recurrence and retreatment, sex-based differences, early-onset disease, and the prognostic and mechanistic significance of the tumour-liver interface.

### Cohort description

#### Study design and setting

KaroLiver is a retrospective, longitudinal, population-based observational cohort at Karolinska University Hospital, Stockholm, Sweden, a national centre for hepatopancreatobiliary surgery. The hospital is the sole provider of elective liver resection for the Stockholm-Gotland healthcare region, which comprises approximately 2.5 million inhabitants. Clinical data are available through the regional electronic health record system, ensuring high data completeness. Vital status and date of death are automatically updated through linkage to the Swedish Population Register, ensuring near-complete ascertainment of overall survival for all patients.

#### Eligibility and recruitment

All consecutive patients who underwent their first curative-intent liver-directed treatment for CRLM at Karolinska University Hospital between February 2012 and January 2022 were included in the current cohort (**Figure 1**). Patients undergoing non-curative-intent procedures or resections of liver metastases from non-colorectal primaries are not included. Re-interventions performed for recurrent CRLM are recorded as additional procedural entries linked to the same patient record, enabling analysis of both primary and repeat liver-directed treatments.

**Figure 1.**
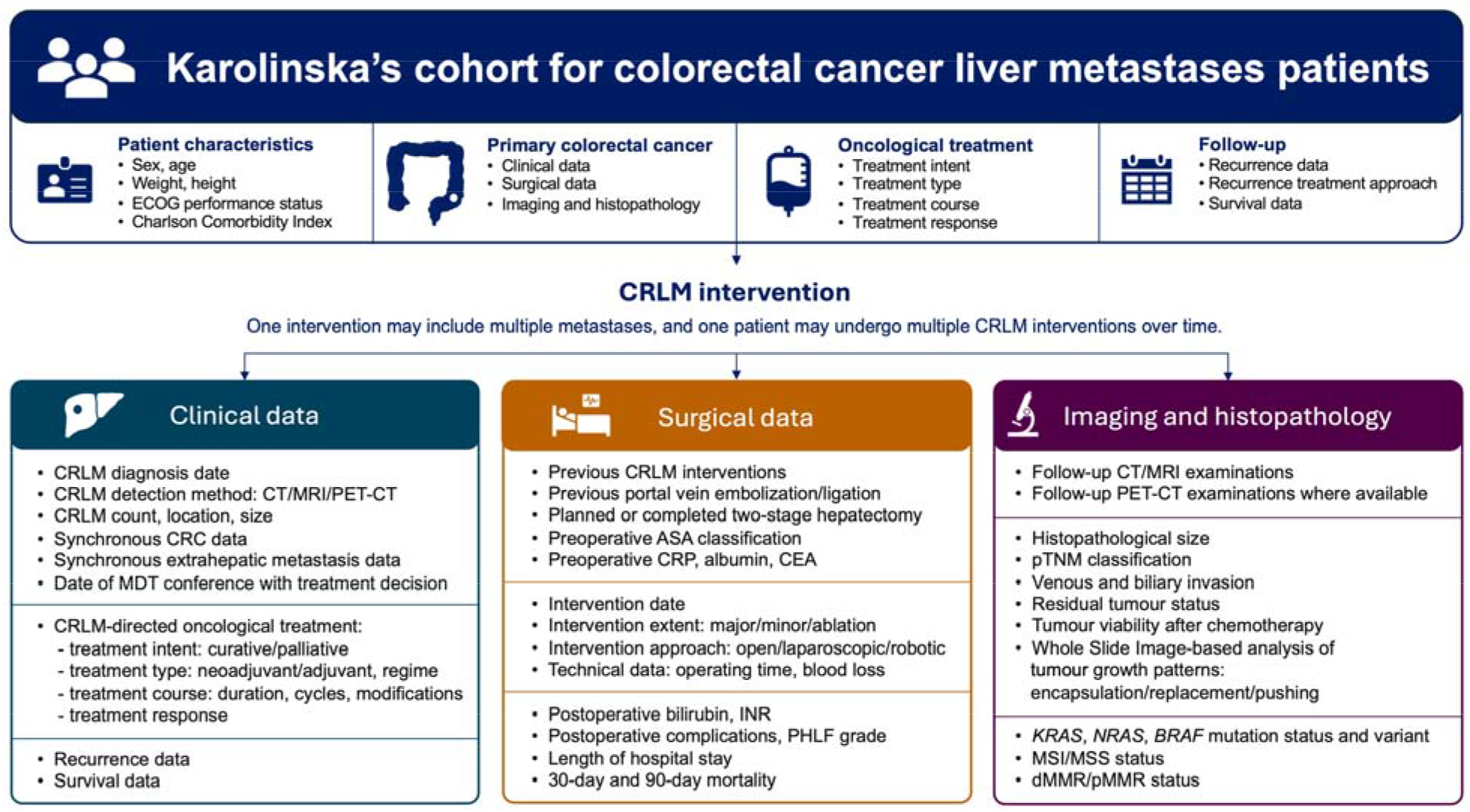
Schematic overview of inclusion criteria and data domains captured in KaroLiver for patients undergoing colorectal cancer liver metastasis-directed interventions at Karolinska University Hospital. ASA, American Society of Anesthesiologists; CEA. carcinoembryonic antigen; CRC, colorectal cancer; CRLM, colorectal liver metastasis; CRP, C-reactive protein; CT, computed tomography; ECOG, Eastern Cooperative Oncology Group; INR, international normalized ratio; MDT, multidisciplinary team; MRI, magnetic resonance imaging; MSI, microsatellite instability; MSS, microsatellite stable; dMMR, deficient mismatch repair; pMMR, proficient mismatch repair; PET-CT, positron emission tomography-computed tomography; PHLF, post-hepatectomy liver failure.

#### Patient and public involvement

No patients or members of the public were involved in the design, conduct, or reporting of this research. Findings will be disseminated through peer-reviewed publication and, where relevant, through patient organisations and institutional communication channels.

#### Multidisciplinary team process

All adult patients with newly diagnosed or recurrent CRLM are discussed at the weekly multidisciplinary team (MDT) liver conference. The MDT consists of HPB surgeons, transplant surgeons, colorectal surgeons, radiologists with specialisation in liver imaging, oncologists, hepatologists, pathologists and clinical nurse specialists.

Prior to the MDT, a standardised imaging evaluation is performed. All patients undergo contrast-enhanced computed tomography (CT) of the chest, abdomen, and pelvis. Magnetic resonance imaging (MRI) of the liver with a liver-specific contrast agent (gadoxetate disodium) is routinely performed. Positron emission tomography-CT (PET-CT) is used selectively, primarily for staging in the context of extrahepatic disease or discordant imaging findings. Dates of imaging and types of examinations are recorded in KaroLiver, and original imaging examinations are archived and linkable through the institutional Picture Archiving and Communication System (PACS).

MDT decisions include the assessment of technical resectability based on the principles of performing complete resection with preservation of a sufficient future liver remnant (FLR) with adequate inflow and outflow, including the need for volume manipulation such as portal vein embolisation (PVE) or hepatic vein embolisation (HVE), recommendation of neoadjuvant chemotherapy, surgical strategy including staged or simultaneous resections in case of synchronously diagnosed CRLM, thermal ablation and non-surgical management. The recommended surgical strategy is generally based on current international and national guidelines [15–17], taking into account patient fitness, comorbidities, and the FLR assessment. The MDT date and decision are recorded for all patients.

#### Surgical procedures and perioperative oncological treatment

All liver interventions are performed electively following MDT discussion. Major hepatectomy is defined as resection of three or more Couinaud segments; trisectionectomy as resection of five or more segments. Minor hepatectomy comprises resection of fewer than three segments, including non-anatomical resections [18,19].

Preoperative CT volumetry is performed routinely to estimate the FLR. An FLR of at least 30% of the total estimated liver volume is generally considered the minimum threshold for safe major resection, depending on liver parenchymal quality and patient factors. When insufficient FLR is anticipated, PVE and/or HVE is performed, with reassessment of FLR after 3 weeks. Associating liver partition and portal vein ligation for staged hepatectomy (ALPPS) is selectively used in patients who require larger oncological resections with insufficient FLR after conventional PVE/HVE.

During an open liver resection, intraoperative ultrasound is routinely employed to verify metastasis location and guide parenchymal-sparing strategies. Parenchymal transection is primarily performed using a Cavitron Ultrasonic Surgical Aspirator (CUSA®), with additional use of blunt-clamp dissection and LigaSure ligation as indicated. Intermittent Pringle manoeuvre, up to 15 minutes of occlusion followed by 5 minutes of releasing, is applied in selected cases. Simultaneous thermal ablation (microwave) is performed under ultrasound guidance when complementary to resection for small (<3 cm) or deep lesions. Minimally invasive hepatectomy (laparoscopic or robotic) is increasingly performed by experienced operators in accordance with institutional criteria. Intraoperative ultrasound and indocyanine green (ICG) fluorescence imaging are routinely used to guide tumour localisation, delineate resection margins, and facilitate parenchyma-sparing resection.

Simultaneous resection is defined as the removal of both the primary tumour and liver metastases during the same general anaesthesia. The decision on simultaneous resection is individualised and based on the extent of liver metastases and the complexity of primary tumour resection. In a reasonably fit patient with limited liver metastases that are resectable by local resection, a simultaneous approach is generally considered, largely irrespective of the complexity of the primary tumour resection. Complex liver resections presuppose uncomplicated resections of the primary tumour. Simultaneous resections were generally initiated with resection of the liver metastases, followed by resection of the primary tumour, the latter performed by a colorectal surgeon. A diverting stoma is considered in cases of high surgical complexity, multiple anastomoses, hypoalbuminemia, poor nutritional status, high frailty or significant intraoperative bleeding, but avoided when possible.

Chemotherapy regimens and decisions regarding neoadjuvant and/or adjuvant treatment generally followed the national recommendations in effect at the time of treatment [15], which were largely based on guidance from the European Society for Medical Oncology (ESMO) (current guidelines, ref. [16]; previous similar guidelines applied throughout the study period). Recommendations on neoadjuvant and/or adjuvant therapy were made by MDTs. Briefly, 5-fluorouracil (5-FU) plus oxaliplatin combination therapies (e.g., FOLFOX) were favoured; 5-FU in combination with irinotecan (e.g. FOLFIRI) was considered on an individual basis or upon progression. If conversion to resectability was the treatment intention, anti-VEGF or anti-EGFR antibodies in combination with chemotherapy were considered; the choice of antibody generally took into account *RAS*/*RAF* mutational status and, increasingly in later years, tumour sidedness; FOLFOXIRI was generally considered for conversion. In metastases with a long disease-free interval, a small solitary lesion, normal CEA, or prior oxaliplatin-based adjuvant therapy, surgery alone was recommended.

#### Postoperative management and follow-up

Following liver intervention, all patients are assessed daily until discharge. Serum bilirubin and INR are routinely measured to assess for post-hepatectomy liver failure (PHLF) according to the International Study Group of Liver Surgery (ISGLS) definition and grading (grades A, B, and C) [20]. Post-hepatectomy haemorrhage and bile leakage are defined and graded according to the respective ISGLS consensus definitions [21,22]. Postoperative complications are graded using the Clavien-Dindo classification [23] and the Comprehensive Complication Index (CCI) [24], both of which are recorded in KaroLiver.

Patients are discharged when clinically stable, tolerating oral intake and with adequate pain management. A standard outpatient follow-up visit is scheduled four to six weeks after surgery. Thereafter, either adjuvant therapy is pursued or structured follow-up is performed at the colorectal surgical unit, typically involving contrast-enhanced CT of the chest, abdomen, and pelvis every 3 to 6 months for the first 2 years, then annually [15]. Serum carcinoembryonic antigen (CEA) is measured at each follow-up visit. Follow-up data, including recurrence dates, sites, and treatment decisions, are systematically retrieved from electronic medical records and recorded in the KaroLiver database.

#### Data collection

Data collection in KaroLiver is motivated by prospectively defined scientific questions. A core component and a specific scientific strength of KaroLiver is the systematic investigation of histological features, including growth patterns defined by the mode of tumour invasion at the tumour-liver interface [6,25], as well as biliary and venous invasion. Nevertheless, the scientific scope of the cohort has expanded, supported by successive ethics amendments, and reflecting the evolution of research questions while remaining grounded in this integrated clinical-histopathological framework.

All patients are identified from records of weekly held multidisciplinary team (MDT) conferences and through institutional databases, ensuring complete, consecutive capture of all treated patients. As KaroLiver is a retrospective observational cohort with consecutive inclusion of all eligible patients, non-participation bias does not apply. Informed consent was waived by the Ethics Review Authority. All data are pseudonymised and stored in a secure, access-controlled database.

The studies related to the cohort are conducted in accordance with the Declaration of Helsinki. Ethics approval for ongoing projects was obtained from the The Swedish Ethical Review Authority (*Etikprövningsmyndigheten*, Dnr 2019-01571, with amendments Dnr 2020-00428, Dnr 2021-06863-02, Dnr 2022-06588-02, and Dnr 2025-05423-02, as well as Dnr 2026-03619-01).

Clinical data are collected in a structured, encrypted database by dedicated physicians with expertise in liver surgery and gastrointestinal oncology, under the supervision of JE and MG. A detailed summary of the collected variables and their definitions is provided in **Table 1**, and **Figure 2** illustrates examples of the various patient trajectories represented in the cohort, depicted using fictitious patient cases to protect personal integrity. No external data sources were used.

**Figure 2.**
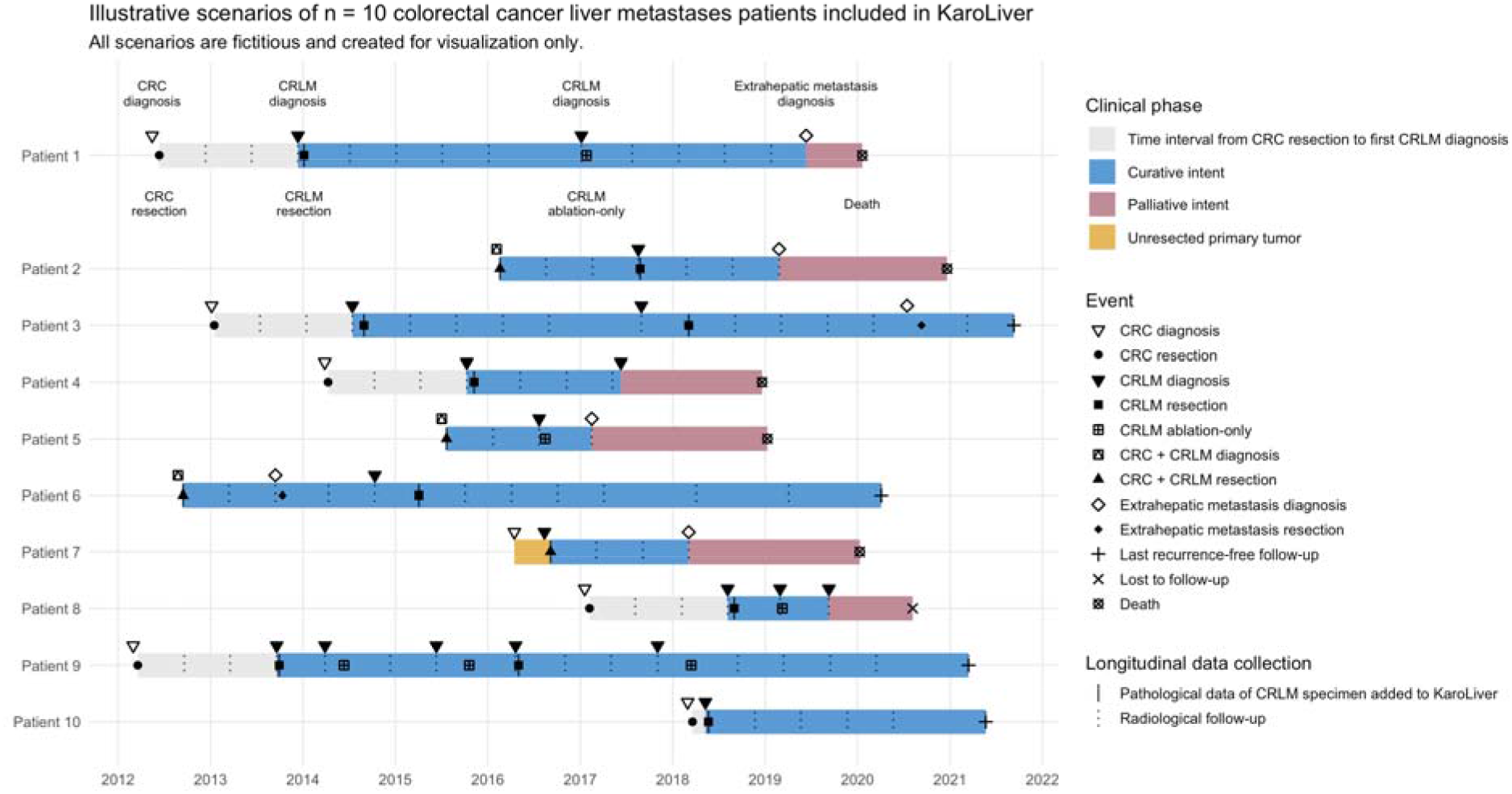
Swimmer plot of fictitious clinical trajectories illustrating the longitudinal data-collection for patients included in KaroLiver. CRC, colorectal cancer; CRLM; colorectal liver metastasis.

**Table 1.** Overview of variable domains and data elements collected in the KaroLiver cohort. ALPPS, associating liver partition and portal vein ligation for staged hepatectomy; ASA, American Society of Anesthesiologists; BMI, body mass index; CCI, Comprehensive Complication Index; CEA, carcinoembryonic antigen; CRC, colorectal cancer; CRLM, colorectal liver metastasis; CRP, C-reactive protein; CT, computed tomography; DPD, dihydropyrimidine dehydrogenase; ECOG, Eastern Cooperative Oncology Group; INR, international normalised ratio; ISGLS, International Study Group of Liver Surgery; MDT, multidisciplinary team; MRI, magnetic resonance imaging; MSI, microsatellite instability; MSS, microsatellite stable; dMMR, deficient mismatch repair; pMMR, proficient mismatch repair; PACS, picture archiving and communication system; PET-CT, positron emission tomography-computed tomography; PHLF, post-hepatectomy liver failure; pT/N/M stage, pathological primary tumour/regional lymph node/distant metastasis stage; R status, residual tumour status; SBRT, stereotactic body radiotherapy.

| Domain | Variables collected |
| --- | --- |
| **Patient identification & demographics** | Database ID, sex, age, weight, height, BMI, ECOG performance status, Charlson Comorbidity Index |
| **Primary colorectal tumour** | Date of CRC diagnosis, tumour location, pT/N/M stage, mucinous histology, vascular/perineural invasion, R status, *KRAS* mutation at primary, oncological treatment, complications after primary resection (Clavien-Dindo, CCI) |
| **CRLM diagnosis** | Date of CRLM diagnosis, age at CRLM diagnosis, date and decisions of MDT meeting, preoperative imaging (CT, MRI, PET-CT with dates), number and size of metastases, lobe distribution (unilobar/bilobar), synchronous vs. metachronous classification, time from CRC to CRLM |
| **Preoperative status** | Preoperative CRP, preoperative albumin, preoperative CEA, ASA classification, portal vein embolisation/ligation, hepatic vein embolisation |
| **Surgical procedure** | Date, type and approach of liver intervention (minimally invasive, (laparoscopic or robotic), open resection, thermal ablation-only), extent of resection (major/minor, operation codes [according to a nationwide Swedish code system]), simultaneous ablation, simultaneous colorectal resection, planned/executed two-stage hepatectomy (including ALPPS), operating time, blood loss, previous liver resection or ablation |
| **Postoperative course** | Postoperative bilirubin and INR (day 5), PHLF grade (ISGLS A/B/C), complications (Clavien-Dindo grade, Comprehensive Complication Index), hospital length of stay, 30- and 90-day mortality |
| **Liver specimen pathology** | Histopathological size of largest metastasis, resection margin (mm), R status, tumour viability after chemotherapy (%), venous and biliary invasion, histopathological growth pattern (encapsulated/replacement/pushing), detailed pathology slide data |
| **Molecular tumour profile** | *KRAS*, *NRAS*, *BRAF* mutation status and specific mutation type (exon, codon), mismatch repair status (MSI/MSS; dMMR/pMMR), tissue source tested (primary vs. metastasis) |
| **Oncological variables, CRLM treatment** | DPD deficiency, neoadjuvant/adjuvant chemotherapy (type, cycles, use of biological agent, radiological response), treatment changes, second-line therapies |
| **Extrahepatic disease** | Presence and date of pulmonary metastases (number, size, bilaterality), |
|  | other extrahepatic disease (location, treatment intent: curative/palliative) |
| **Recurrence and retreatment** | Date of first recurrence, organ/site of recurrence (liver, lung, peritoneum, other), indicator for recurrence detection (radiology/clinical/CEA), curative-intent vs. palliative retreatment (type, date), reasons for non-curative management, dates and type of subsequent liver or extrahepatic interventions (resection, thermal ablation, SBRT, other) |
| **Survival** | Date of death, cancer-related vs. other cause of death, date last known alive, overall survival, recurrence-free survival |
| **Imaging archive** | Linked CT/MRI examinations from CRLM diagnosis and at each recurrence, accessible through hospital PACS system |

Key domains include: patient demographics and comorbidity; primary CRC characteristics (tumour location, stage, molecular profile); CRLM characteristics at time of liver intervention (number, size, distribution, synchronous vs. metachronous); preoperative workup; surgical procedure; postoperative outcomes; liver specimen pathology (including histopathological growth pattern, margin status, tumour viability, and venous invasion); molecular tumour profile (*RAS* and *BRAF* mutations, mismatch repair status); perioperative oncological treatment; recurrence data (date, site, treatment intent); and survival (date of death or last known alive, cause of death). Additionally, all original CT and MRI examinations from the time of CRLM diagnosis and from recurrence imaging are archived and can be linked to the KaroLiver database via the hospital PACS system.

The descriptive statistics and analyses of recurrence-free survival and overall survival for this manuscript were performed using R version 4.5.3 (R Foundation for Statistical Computing, Vienna, Austria) in RStudio version 2026.01.1+403 (Posit Software, PBC, Boston, MA, USA). Missing data are presented as proportions for each variable and were not imputed. No assumptions were made regarding the nature of missingness. Handling of incomplete follow-up is adapted to the specific research question and outcome under study and is defined and reported transparently in each individual analysis.

#### Characteristics of study participants

Between February 2012 and January 2022, a total of 811 patients underwent their first curative-intent liver-directed treatment for CRLM at Karolinska University Hospital and were enrolled. These patients underwent a total of 1204 procedures (811 first interventions and 393 re-interventions). Baseline characteristics of all patients, related to their first liver intervention, are presented in **Table 2**.

**Table 2.** Baseline characteristics of patients in the KaroLiver cohort undergoing their first CRLM-directed intervention at Karolinska University Hospital. Values are n (% of all patients) or median (interquartile range, IQR) unless otherwise stated. ASA, American Society of Anesthesiologists; CEA, carcinoembryonic antigen; CI, confidence interval; CRC, colorectal cancer; CRLM, colorectal liver metastasis; ISGLS, International Study Group of Liver Surgery; pN stage, pathological regional lymph node stage; pT stage, pathological primary tumour stage.

| Characteristics | All patients, N = 811 | Missing, n (%) |
| --- | --- | --- |
| **Demographics** |  |  |
| Sex, female, n (%) | 307 (37.9%) | 0 |
| Age at first liver intervention, years, median (IQR) | 67 (59–74) | 0 |
| Body mass index at first liver intervention, kg/m<sup>2</sup>, median (IQR) | 25.3 (22.9–28.0) | 42 (5.2%) |
| ASA classification at first liver intervention, n (%) | 810 (99.9%) | 1 (0.1%) |
| I–II | 427 (52.7%) |  |
| III–IV | 383 (47.2%) |  |
| Charlson Comorbidity Index at first CRLM diagnosis, n (%) | 810 (99.9%) | 1 (0.1%) |
| 6–7 | 189 (23.3%) |  |
| 8–9 | 442 (54.5%) |  |
| ≥10 | 179 (22.1%) |  |
| **Primary colorectal tumour** |  |  |
| Primary tumour location, n (%) | 804 (99.1%) | 7 (0.9%) |
| Right colon (caecum – transverse colon) | 208 (25.6%) |  |
| Left colon (splenic flexure – sigmoid) | 294 (36.3%) |  |
| Rectum | 302 (37.2%) |  |
| Resection of primary tumour (yes) | 749 (92.4%) | 14 (1.7%) |
| pT stage, n (%) | 729 (89.9%) | 82 (10.1%) |
| T0 | 12 (1.5%) |  |
| T1–T2 | 93 (11.5%) |  |
| T3–T4 | 624 (76.9%) |  |
| pN stage, n (%) | 728 (89.8%) | 83 (10.2%) |
| N0 | 227 (28.0%) |  |
| N1–N2 | 501 (61.8%) |  |
| **Metastatic disease at time of first liver intervention** |  |  |
| Synchronous CRLM (diagnosed ≤ 3 months from CRC), n (%) | 478 (58.9%) | 0 |
| Number of liver metastases, median (IQR) | 2 (1–4) | 0 |
| 1, n (%) | 309 (38.1%) |  |
| 2–3, n (%) | 251 (30.9%) |  |
| ≥ 4, n (%) | 251 (30.9%) | |
| Size of largest metastasis, mm, median (IQR) (preoperative radiological or intraoperative ultrasound measurement) | 25 (15–40) | 5 (0.6%) |
| Bilobar disease, n (%) | 317 (39.1%) | 2 (0.2%) |
| Extrahepatic disease at time of first liver intervention, n (%) | 135 (16.6%) | 1 (0.1%) |
| CEA at liver intervention, $\mu\text{g/L}$, median (IQR) | 5.7 (2.9–19.0) | 442 (54.5%) |
| **Molecular tumour characteristics** |  |  |
| *RAS* mutation (*KRAS* or *NRAS*), n (%) | 191 (23.6%) | 436 (53.8%) |
| *BRAF V600E* mutation, n (%) | 27 (3.3%) | 459 (56.6%) |
| Microsatellite instability, n (%) | 12 (1.5%) | 621 (76.6%) |
| **Perioperative oncological treatment** |  |  |
| Neoadjuvant chemotherapy, n (%) | 502 (61.9%) | 10 (1.2%) |
| Adjuvant chemotherapy, n (%) | 386 (47.6%) | 85 (10.5%) |
| **Liver intervention** |  |  |
| Major hepatectomy (≥ 3 segments), n (%) | 330 (40.7%) | 0 |
| Right hemihepatectomy, n (%) | 238 (29.3%) |  |
| Left hemihepatectomy, n (%) | 28 (3.5%) |  |
| Trisectionectomy, n (%) | 5 (0.6%) |  |
| Minor hepatectomy (< 3 segments), n (%) | 452 (55.7%) | 0 |
| Thermal ablation only, n (%) | 29 (3.6%) | 0 |
| Simultaneous ablation, n (%) | 92 (11.3%) | 0 |
| Simultaneous CRC resection, n (%) | 138 (17.0%) | 0 |
| Operating time, minutes, median (IQR) | 211 (150–286) | 397 (49.0%) |
| Intraoperative blood loss, mL, median (IQR) | 400 (200–800) | 174 (21.5%) |
| pR0 resection (clear margins ≥ 1 mm), n (%) | 491 (60.5%) | 62 (7.6%) |
| **Postoperative outcomes** |  |  |
| Highest Clavien-Dindo grade, n (%) | 810 (99.9%) | 1 (0.1%) |
| 0 (no complication) | 351 (43.3%) |  |
| I–II (minor) | 295 (36.4%) |  |
| ≥ IIIa (major) | 164 (20.2%) | |
| Post-hepatectomy liver failure ISGLS grade B or C, n (%) | 33 (4.1%) | 17 (2.1%) |
| Length of hospital stay, days, median (IQR) | 8 (6–11) | 12 (1.5%) |
| 90-day mortality, n (%) | 23 (2.8%) | 0 |
| **Recurrence and survival** |  |  |
| Any recurrence, n (%) | 548 (67.6%) | 0 |
| Site of first recurrence, n (% of patients with recurrence) |  |  |
| Liver only | 225 (41.1%) |  |
| Extrahepatic only | 215 (39.2%) |  |
| Combined liver and extrahepatic | 108 (19.7%) |  |
| Curative-intent retreatment after recurrence, n (% of patients with recurrence) | 277 (50.5%) | 29 (5.3%) |
| Recurrence-free survival, months, median (95% CI) | 10.4 months (95% CI 9.2–11.9 months) | 0 |
| Overall survival, months, median (95% CI) | 51.0 months (95% CI 46.2–57.1 months) | 0 |
| 5-year overall survival, % (95% CI) | 44.9% (95% CI 41.2–48.8%) | 0 |

Median age at first liver intervention was 67 years (IQR 59–74), and 37.9% of patients were female. Synchronous CRLM, defined as diagnosis of the CRLM within 3 months of the primary colorectal cancer, was present in 58.9% of patients. The median number of liver metastases at the time of intervention was 2 (IQR 1–4), and the median size of the largest metastasis was 25 mm (IQR 15– 40 mm).

Major hepatectomy (≥3 liver segments) was performed in 40.7% of patients, minor hepatectomy (<3 segments) in 55.7%, and ablation only in 3.6% as the first liver intervention. Among evaluable first-intervention cases, a pR0 resection margin (≥1 mm) was achieved in 65.6%. Neoadjuvant chemotherapy was administered to 61.9% of patients and adjuvant chemotherapy to 47.6% after liver intervention. Molecular tumour profiling included *RAS* mutation status in 46.2% of cases, *BRAF V600E* status in 43.4%, and MSI status in 23.4%.

Follow-up time was defined as the interval from the first CRLM-directed intervention to the date of death or the date on which the patient was last known to be alive. Using the reverse Kaplan–Meier method, the median follow-up time was 75.8 months (95% CI 69.3–81.9 months), corresponding to 6.3 years (95% CI 5.8–6.8 years). At the end of follow-up, 357 patients were alive (44.0%) and 454 patients had died (56.0%). Perioperative mortality was low, with 30-day and 90-day rates of 0.7% and 2.8%, respectively. Median recurrence-free survival (RFS), defined as the time from the first liver intervention to recurrence or death, whichever came first, was 10.4 months (95% CI 9.2–11.9 months), median overall survival (OS) was 51.0 months (95% CI 46.2–57.1 months), and 5-year overall survival was 44.9% (95% CI 41.2–48.8%) (**Figure 3**).

**Figure 3.**
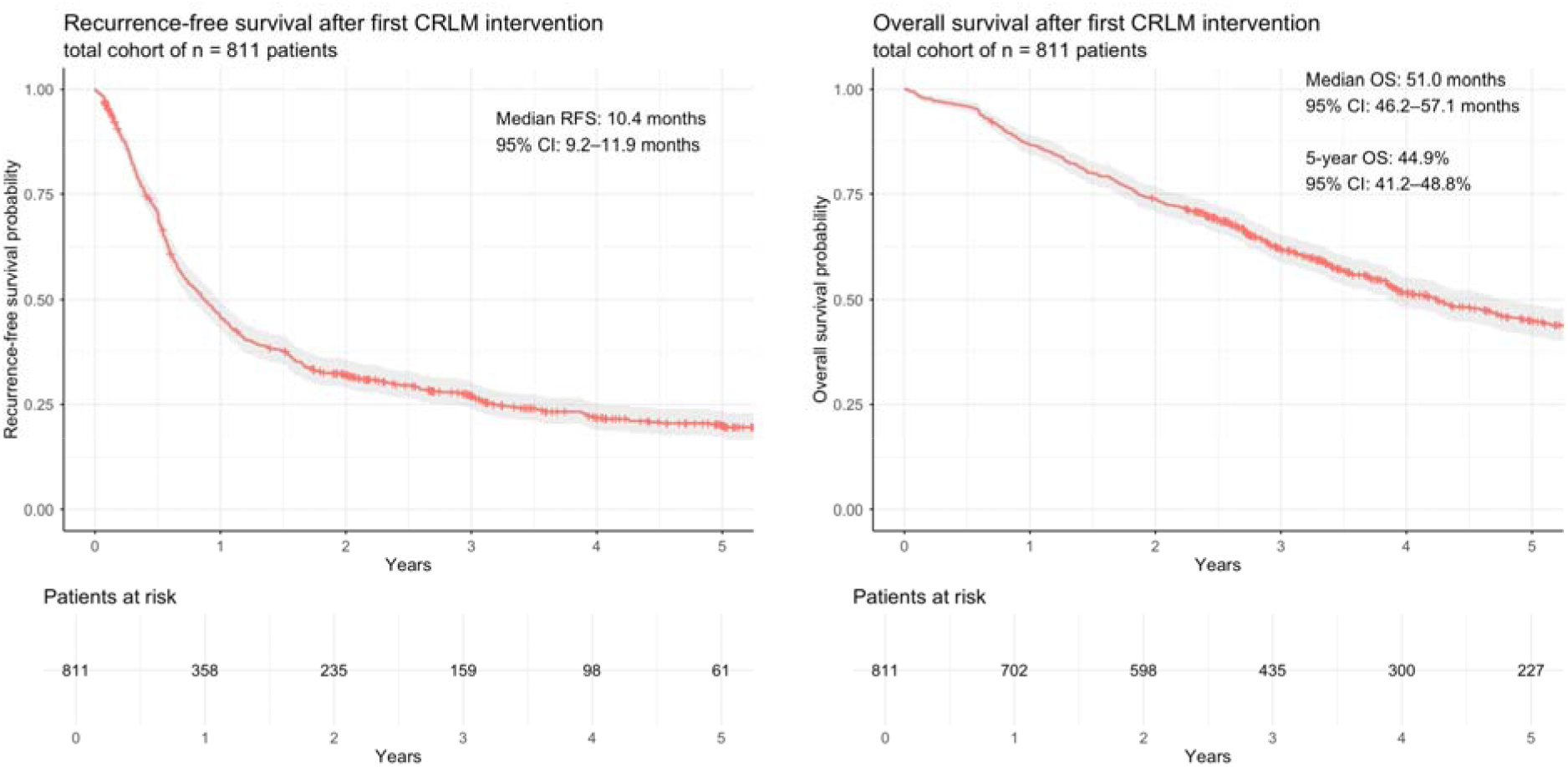
Kaplan-Meier curves of recurrence-free and overall survival of patients in the KaroLiver cohort after the first colorectal liver metastasis intervention. CRLM, colorectal liver metastasis; RFS, recurrence-free survival; OS, overall survival.

#### Histology

A key feature of KaroLiver is the systematic and comprehensive capture of histopathological data from surgical resection specimens, as described in detail previously [5]. All liver resection specimens are processed according to a standardised sampling protocol developed at the Department of Clinical Pathology and Cancer Diagnostics, Karolinska University Hospital Huddinge, which approximates a panoramic section through the largest cross-sectional diameter of each metastasis to ensure representative coverage of the entire tumour-liver invasion front for all tumours. Diagnostic slides are scanned as whole-slide images (WSIs) and linked to KaroLiver’s clinical dimensions. Growth patterns are assessed using international consensus criteria [6] and classified as encapsulated (i.e., “desmoplastic”), replacement, or pushing. For n = 273 operations, the patterns have already been scored using WSI-based manual digital annotation across the entire invasion front under the supervision of a specialized HPB pathologist (CFM), enabling quantitative, fine-grained analyses of growth pattern fractions [5]. Additional histopathological variables include resection margin (mm), pR0/R1 status, tumour viability after preoperative chemotherapy (%), and venous and biliary invasion extracted from pathology reports. Where available, immunohistochemistry from routine pathological work-up is collected and digitised for quantitative integration with clinical data. So far, WSIs from n = 273 resections, corresponding to 562 tumours and 1292 WSIs, have been annotated for growth patterns along the entire invasion front (**Table 3**), and additional pathological data have been extracted from the pathology reports.

**Table 3.** Histopathological data in the KaroLiver cohort for all CRLM specimens, surgically resected at Karolinska University Hospital from February 2012 onwards. Values are n (% of all resections) or median (interquartile range, IQR) unless otherwise stated. ^1^viable tumour only included for patients after neoadjuvant treatment.

| Pathological variable (liver specimen) | All resections, N = 1017 | Missing, n = % |
| --- | --- | --- |
| Histopathological size, largest metastasis, mm, median (IQR) | 22 (13 – 37) | 108 (10.6%) |
| Manually annotated growth pattern of the entire invasion front, n (%) | 273 (26.8%) | 744 (73.2%) |
| Predominantly replacement growth pattern ($\geq 50\%$), n (% of patients with available growth pattern) | 129 (47.3%) | |
| Individual tumours assessed for growth pattern analysis, n | 562 |  |
| Slides scanned and assessed for growth pattern analysis, n | 1292 |  |
| Venous invasion, n (%) | 455 (44.7%) | 179 (17.6%) |
| Biliary invasion, n (%) | 205 (20.2%) | 179 (17.6%) |
| Tumour viability after chemotherapy, % in the nodule with highest viability, median (IQR) <sup>1</sup> | 50 (30–70) | 458 (45.0%) |

Archived paraffin-embedded tissue blocks are retained routinely, enabling future immunohistochemical or molecular analyses. Biobank-stored serum and plasma samples from Stockholm’s Medical Biobank, collected with patient consent, can principally be linked to the cohort for biomarker analyses and accessed for defined research questions.

#### Findings to date

The main translational study from KaroLiver so far characterised the fibrotic tissue encapsulating “desmoplastic” CRLM [5]. Using KaroLiver specimens, the study linked improved RFS and OS to increasing tumour encapsulation and decreasing tumour-hepatocyte contact. Growth patterns were scored in a subset of n = 231 tumours included in KaroLiver (2012 – 2015) by WSI-based digital annotations across the entire tumour-liver invasion front, revealing that partially encapsulated tumours form a distinct intermediate prognostic group. Immunostaining revealed a zonated capsular structure, which, together with the clinical data implied that encapsulation develops where tumour invasion into the liver parenchyma is impaired, representing a reparative process that opposes aggressive tumour invasion; these findings led to the proposal to rename the “desmoplastic” pattern to “encapsulated”, which more closely reflects this biology [26]. Building on this, additional studies investigated the specific impact of PHLF on oncological recurrence following major hepatectomy for CRLM [27], and examined sex-associated aspects of referral patterns, surgical resectability, and survival [11].

#### Strengths and limitations

KaroLiver has several key strengths. First, the cohort is consecutive and population-based, capturing all patients undergoing curative-intent liver-directed treatment at a high-volume tertiary referral centre serving a large healthcare region covering urban and suburban areas and, in part, rural areas on the outskirts of Stockholm. This minimises selection bias and supports generalisability to comparable European HPB centres.

A second strength is the systematic and granular recording of recurrence data. This enables detailed analyses of disease trajectory, retreatment patterns, and survival beyond first hepatectomy, which is not available in most published CRLM cohorts.

Third, the combination of linked CT/MRI examinations from CRLM diagnosis and at each recurrence, comprehensive liver specimen pathology data, including histopathological growth pattern, and molecular tumour profiling (*RAS*, *BRAF*, mismatch repair status), provides a strong platform for translational and radiology-based research.

Fourth, the cohort’s size and continuous accrual over more than a decade enable subgroup analyses with relatively high statistical power, including time-period trends in surgical techniques and oncological management.

Several limitations exist. The observational design inherently limits causal inference, and the potential for residual confounding must be considered. Treatment strategies and follow-up protocols evolved over the study period, introducing potential calendar-period confounding; this can be addressed in analyses by adjusting for the year of surgery or by conducting sub-period analyses. Molecular tumour profiling data are not available for all patients, particularly those treated in the earlier years of the cohort, reflecting the gradual adoption of routine molecular testing in clinical practice. Finally, the cohort is limited to a single institution, which, while ensuring data quality and completeness, may limit generalisability to centres with different patient populations or surgical volumes.

Researchers planning similar cohorts should consider two practical lessons from the KaroLiver experience. First, data storage is a critical component: the use of a structured database platform with defined entry fields and built-in range checks reduces the risk of data loss, entry errors, and inconsistencies that can compromise data quality over time. Second, careful consideration should be given to the scope of variables collected. Including sufficient variables to address the intended research questions is essential, but an overly broad variable list risks becoming unmanageable, increases the burden on data collectors, and may compromise data completeness and consistency. Clear, pre-specified variable definitions agreed upon before data collection begins are fundamental to ensuring reproducibility and comparability across time and between data entry personnel.

### Collaboration

The KaroLiver cohort aims to serve as an open research platform, and the research group actively encourages collaboration with other clinical research groups and translational scientists. Researchers interested in using the KaroLiver data for specific research questions are encouraged to contact the principal investigators (JE and MG) to discuss collaboration opportunities.

Access to de-identified data can be arranged upon approval of a study proposal by the principal investigators, confirmation of all applicable ethical approvals from all relevant parties, and completion of any relevant data/material transfer agreements and/or research collaboration agreements, as well as any further regulatory and legal documents required for the specific project. Restrictions may apply in accordance with applicable data protection and research ethics legislation and institutional policies.

#### Future plans

The KaroLiver cohort plans to continue accruing patients and longitudinal outcome data. Several research streams are planned or ongoing, broadly spanning tumour biology and pathology, surgical and locoregional treatment outcomes, recurrence and retreatment, patient-related factors, and health equity.

Near-term research priorities include the evaluation of sex-based differences in recurrence patterns, retreatment strategies, and survival; characterisation of early-onset versus average-onset CRLM with respect to metastatic burden, molecular profile, surgical management, and oncological outcomes; and the impact of follow-up intensity on recurrence detection and survival. Studies leveraging the linked imaging archive and histopathological growth pattern data are also planned. Serum and plasma proteomics from biobank-linked samples will be used to investigate circulating correlates of histological features such as growth patterns.

External validation of established prognostic scores, including and excluding histological variables, for recurrence and survival is ongoing.

Follow-up data collection and database updates will continue in accordance with ethical approvals. Long-term OS and RFS data are expected to mature further over the coming years, allowing robust 10-year outcome analyses.

## Supporting information

Supplemental Table 1: STROBE Checklist

## Data Availability

All data produced in the present study are available upon reasonable request to the authors. Data sharing may be arranged for approved research collaborations upon mutual ethical approval from all relevant parties, signature of an appropriate data access and/or research collaboration agreement, and, where applicable, regulatory clearance. De-identified datasets will be made available to qualified researchers for approved analyses.

## Author’s Contributions

MG, JE, and CFM conceptualised KaroLiver and supervised data collection. ES contributed expertise on research project design. SH, YH, ETQ, MSS, LB, and AKH collected and curated pathological information and WSIs, and RB collected pathological information. BB interpreted pathological data and devised pathological sampling procedures. AV, CL, and NG curated and analysed clinical and pathological data, and integrated diverse data sources. JS collected and interpreted clinical data.

The authors used ChatGPT (OpenAI; mainly version GPT-5.5 Thinking; accessed May and June 2026) for code troubleshooting and refinement of the code used to derive variables in Tables 2 and 3, and to derive visual presentation options for Figure 1. Figure 1 was generated in PowerPoint (Microsoft) by the authors; selected graphical elements were created using royalty-free icons from the Microsoft 365 stock content library.

Claude (Anthropic; Claude Sonnet 4.6; accessed May and June 2026) was used for language editing and refinement of selected manuscript text. The authors reviewed, edited and verified all AI-assisted code and outputs, and take full responsibility for the accuracy and integrity of the final manuscript.

## Ethics approval

The study was approved by The Swedish Ethical Review Authority (Etikprövningsmyndigheten, Dnr 2019-01571), with successive amendments (Dnr 2020-00428, Dnr 2021-06863-02, Dnr 2022-06588-02, Dnr 2025-05423-02, and 2026-03619-01). The study was conducted in accordance with the Declaration of Helsinki. Individual informed consent was waived by the Ethics Review Authority.

## Data sharing statement

Data sharing may be arranged for approved research collaborations upon mutual ethical approval from all relevant parties, signature of an appropriate data access and/or research collaboration agreement, and, where applicable, regulatory clearance. De-identified datasets will be made available to qualified researchers for approved analyses. Researchers interested in accessing the KaroLiver data or linked imaging/pathology materials are encouraged to contact the corresponding authors to discuss collaboration and data-sharing conditions.

